# Strategic anti-SARS-CoV-2 serology testing in a low prevalence pandemic: The COVID-19 Contact (CoCo) Study in health care professionals

**DOI:** 10.1101/2020.08.06.20169250

**Authors:** Georg Behrens, Anne Cossmann, Metodi V. Stankov, Bianca Schulte, Hendrik Streeck, Reinhold Förster, Berislav Bosnjak, Stefanie Willenzon, Anna-Lena Boeck, Anh Thu Tran, Thea Thiele, Theresa Graalmann, Moritz Z. Kayser, Anna Zychlinsky Scharff, Christian Dopfer, Alexander Horke, Isabell Pink, Torsten Witte, Martin Wetzke, Diana Ernst, Alexandra Jablonka, Christine Happle

## Abstract

**Background:** Serology testing is explored for epidemiological research and to inform individuals after suspected infection. During the COVID-19 pandemic, frontline healthcare professionals (HCP) may be at particular risk for infection. No longitudinal data on functional seroconversion in HCP in regions with low COVID-19 prevalence and low pre-test probability exist.

**Methods:** In a large German university hospital, we performed weekly questionnaire assessments and anti-SARS-CoV-2 IgG measurements with various commercial tests, a novel surrogate virus neutralization test, and a neutralization assay using live SARS-CoV-2.

**Results:** From baseline to week six, n=1,080 screening measurements for anti-SARS CoV-2 (S1) IgG from n=217 frontline HCP (65% female) were performed. Overall, 75.6% of HCP reported at least one symptom of respiratory infection. Self-perceived infection probability declined over time (from mean 20.1% at baseline to 12·4 % in week six, p<0.001). In sera of convalescent PCR-confirmed COVID-19 patients, we measured high anti-SARS-CoV-2 IgG levels, obtained highly concordant results from ELISAs using e.g. the S1 spike protein domain and the nucleocapsid protein (NCP) as targets, and confirmed antiviral neutralization. However, in HCP the cumulative incidence for anti-SARS-CoV-2 (S1) IgG was 1.86% for positive and 0.93% for equivocal positive results over the six week study period. Except for one HCP, none of the eight initial positive results were confirmed by alternative serology tests or showed in vitro neutralization against live SARS CoV-2. The only true seroconversion occurred without symptoms and mounted strong functional humoral immunity. Thus, the confirmed cumulative incidence for neutralizing anti-SARS-CoV-2 IgG was 0.47%.

**Conclusion:** When assessing anti-SARS-CoV-2 immune status in individuals with low pre-test probability, we suggest confirming positive results from single measurements by alternative serology tests or functional assays. Our data highlight the need for a methodical serology screening approach in regions with low SARS-CoV-2 infection rates.

## Background

Uncertain rates of asymptomatic infections have raised concerns about a potentially high rate of undiagnosed infections with Severe Acute Respiratory Syndrome Coronavirus 2 (SARS-CoV-2) ^1,2^. Health Care Professionals (HCP) were shown to be at risk of infection during previous coronavirus outbreaks ^3,4^. During the current pandemic, asymptomatic SARS-CoV-2 infection ^5^ and onward transmission of SARS-CoV-2 in HCP has been demonstrated ^6,7^. However, nosocomial spread to HCP depend on regional infection patterns ^1,8^. In Wuhan, where the SARS-CoV-2 outbreak was first reported, the incidence of COVID-19 was higher in HCP than the general public ^9^. In contrast, studies from Spain and Belgium demonstrated SARS-CoV-2 infection rates of 6 to 30% irrespective of patient contact ^8,10^, likely reflecting pandemic spread in the general population. Thus, both local infection dynamics and work place precautions against SARS-CoV-2 transmission such as personal protection equipment (PPE) affect an HCP’s risk of becoming infected.

SARS-CoV-2 specific B cell responses typically lead to detectable antibody titers and fully positive rates at about 18 days after the initial onset of symptoms ^11^. Seroepidemiological studies can help to catalog those who have been previously infected (including mild or subclinical infections) and may help identify at-risk populations ^12^. Longitudinal analysis of humoral immunity is particularly valuable in persons at high risk for exposure such as HCP. We ^13^ and others ^14^ have demonstrated that the degree of humoral immune responses as assessed by ELISA correlates with severity of COVID-19. Consequently, it is important to explore whether asymptomatic SARS-CoV-2 infections also lead to detectable and functional antibody responses. Various in-house and commercial serological testing systems for SARS-CoV-2 specific immunoglobulins (Ig) to support clinical decision making and epidemiological studies are currently on the market or being developed ^15^. However, interpretation of individual anti-SARS-CoV-2 serology results in HCP and others depends not only on sensitivity and specificity of the testing systems, but also on the regional prevalence of SARS-CoV-2 infections and the resulting pre-test probability of disease.

To further assess the validity of different serological testing systems in frontline HCP we carried out the prospective COVID-19 Contact (CoCo) Study at Hanover Medical School, a large university hospital in Northern Germany. Our aims were: 1) to obtain longitudinal data about the actual and self-perceived risk of infection, 2) to detect clinically silent seroconversions, 3) to assess the performance of serological testing systems (ELISAs and rapid test) detecting SARS-CoV-2 antibodies (total, IgG, IgA), and 4) to determine the quality of systemic humoral immune responses detected by ELISAs by employing a novel *in vitro* virus inhibition test and neutralisation assay using live virus.

## Methods

### Study design, enrolment, and follow up

The CoCo study ^13^ is an ongoing prospective study which longitudinally monitors SARS-CoV-2-specific IgG serum levels as well as symptoms of respiratory infection, work environment, and self-perceived risk. The study (DRKS00021152) is approved by local authorities (Data Security Management and Institutional Review Board of Hanover Medical School, approval #8973_BO_K_2020). Study participants in the CoCo cohort 1.0 (Figure 1) were enrolled between March 23rd and April 17^th^ 2020. After written informed consent was obtained, participants were asked to provide blood specimens weekly during the first two months, followed by monthly testing. To assess the self-perceived probability of having already contracted SARS-CoV-2, the following question was asked at each visit: “How high do you rate the probability of having been infected so far? (0-100%)”. Here, we report on the first six weeks of the CoCo 1.0 cohort.

**Figure 1:**
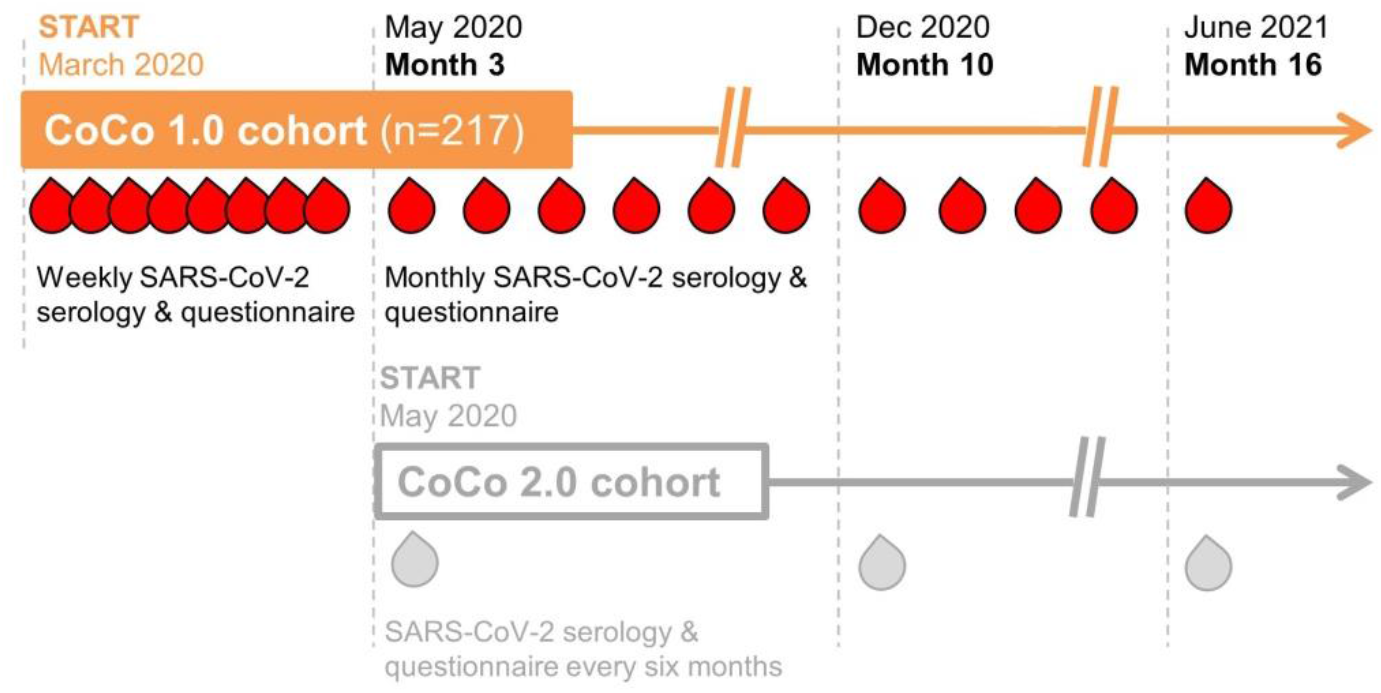
Design of the CoCo Study. The CoCo cohort 1.0 comprises n=217 frontline HCP from emergency departments, infectious and pulmonary disease inpatient units, ICUs, pediatric departments and other units involved in COVID-19 patient care for weekly serologic screening for SARS-CoV-2 during the first two months followed by monthly testing. CoCo cohort 2.0 enrollment started in May 2020 to recruit additional n=1,000 HCP from other clinical departments of Hanover Medical School for serologic assessments every six months.

### Laboratory testing

We used a semiquantitative ELISA for IgG based on the SARS-CoV-2 S1 spike protein domain/receptor binding domain (Euroimmun, Lübeck, Germany) for primary testing. For additional secondary analyses in all baseline samples, positive controls, and positive or equivocal positive sera, an anti-SARS-CoV-2 S1 IgA, an anti-SARS-CoV-2 nucleocapsid protein (NCP) IgG ELISA (Euroimmun, Lübeck, Germany), and a WANTAI SARS-CoV-2 antibody rapid test (SZABO SCANDIC, Vienna, Austria - CE) was used (for more details, see *Suppl. Information*).

The neutralisation assay^16^ was performed using an *in vitro*-propagated SARS-CoV-2 strain isolated in Bonn, Germany via nasopharyngeal swabbing of a patient from Heinsberg,Germany. Briefly, to test SARS-CoV-2 neutralisation capacities, neutralising titers were calculated as the reciprocal of serum dilutions resulting in neutralisation of 50% or 90% input virus (NT50/NT90, respectively), read out as reduction in the number of plaques (for more details, see *Suppl. Information*). The surrogate virus neutralisation test (sVNT) will be described elsewhere in detail ^17^ and is based on the hypothesis that virus neutralising antibodies also interfere with the binding of the receptor-binding domain (RBD) of SARS-CoV-2 to surface-immobilised ACE2. In brief, hACE2 protein (Trenyzme) was coated at 300 mM on Nunc-Immuno plates (Thermo Scientific) and then blocked with 2% bovine serum albumin (BSA, Sigma) and 0.1% Tween. 6ng/ml His-tag-conjugated SARS-CoV-2-S-RBD (Trenzyme) was pre-incubated (or not) with sera at different concentrations for 1 h at 37°C and then added for 1.5 hrs to the ACE2-coated plates. Unbound SARS-CoV-2-S-RBD was washed off before anti-His peroxidase-labelled mAb (Clone 3D5) was added for 1h at 37°C. After final washing, colorimetric signal was developed by adding, 3,3’,5,5’-tetramethylbenzidine (Sigma) and stopped by adding H_2_SO_4_. Absorbance at 450 nm and 570 nm were acquired using a SpectraMax ID3 microplate reader (Molecular Devices). Inhibition (%) was calculated as (1 - Sample OD value/Average SARS-CoV-2-S-RBD OD value) x100.

### Statistical analysis

Data were analysed using SPSS^®^ Statistics (Version 26) and GraphPad Prism^®^ (Version 5). Data are presented as mean + SEM or median and range. For statistical evaluation, Pearson correlation or Fisher’s exact test was performed and differences between groups were assessed by t-test or ANOVA with post hoc Kruskal-Wallis testing when more than two groups were compared.

## Results

Figure 1 shows the design of the CoCo study. The CoCo 1.0 cohort study follows n=217 HCP (65% female) from units involved in COVID-19 patient care with longitudinal collection of biomaterials and questionnaire-based information on health status and working and living conditions. From baseline to week six, for a total of n=1,080 anti-SARS CoV-2 IgG measurements were performed. Follow-up rates were high, with 79·8% of possible timepoints collected (mean 4·98 timepoints per HCP, range 1-7 per HCP). Of all HCP, 29·0% reported respiratory symptoms during the two weeks before the first (baseline) timepoint with higher frequencies in men vs. women (39·5% vs. 23·4%, Fisher Exact: 6·3, p=0·01). The presence of children below the age of 12 years in the same household was associated with a higher rate of respiratory infections (25.6% in childless households vs. 43·9% for HCP sharing a household with children under 12 years, Fisher Exact: 6·1, p=0.018). 8·8% of study participants reported being on sick leave and 3·3% reported having been quarantined during the four weeks prior to enrollment.

At baseline, 45·2% of HCP reported at least one symptom suggestive of respiratory illness, but this rate declined gradually to 25·4% by week six. Over the full study period, 75·6% of HCP reported at least one respiratory symptom. The rates for sick leave and quarantine over the full six-week period were 2·8% and 2·3%, respectively. Upon enrollment, 16·1% of HCP reported having been in direct contact with a confirmed SARS-CoV-2 infected person. During the six week period study, the cumulative proportion of HCP reporting contact with confirmed infected persons was 30%.

During this time, only 3·2% of all CoCo 1.0 cohort participants were tested for SARS-CoV-2 by PCR from nasopharyngeal swabs, and all tests yielded negative results. The mean self-perceived infection probability decreased from a mean of 20·9% upon enrollment to 12·5% at week six(p<0·001, Figure 2). This decline was evident in men and women, with women rating their risk higher than men (males: decline from 15·4% to 8·1% vs. females decline from 24·1% to 14·8%).

**Figure 2:**
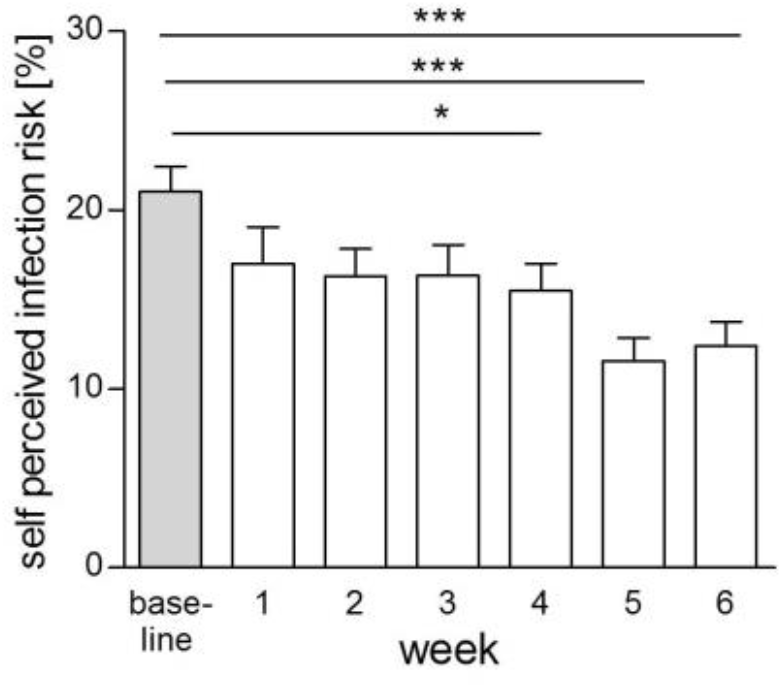
Self-perceived infection risk over time. Reduction of mean self-perceived infection risk of all CoCo cohort 1.0 participants answering this question over the first six weeks. Bars display mean + SEM, *p<0·05, *** p<0·001.

As previously reported, the baseline prevalences for anti-SARS-CoV-2 S1 IgG and IgA of the CoCo 1.0 cohort were low, at 0·9-1·8% and 4·1-8·7%, respectively ^13^, and no cases of COVID-19 were observed in study participants until week six of observation. To assess the concordance of various testing systems in a low prevalence setting, we performed additional tests for anti-SARS-CoV-2 NCP IgG, as well as a SARS-CoV-2 antibody rapid test on the same set of samples. First, we assessed the sensitivity and specificity for all assays in convalescent PCR^+^ COVID-19 patients. The concordance of testing results was highest between the anti-SARS-CoV-2 S1 and anti-SARS-CoV-2 NCP IgG ELISAs (92·7%, Suppl. Fig. 1). The measured levels of anti-SARS-CoV-2 S1 IgG and anti-SARS-CoV-2 NCP IgG results also correlated closely (Suppl. figure 2A). However, their combined sensitivity within the control cohort was only 87·5% (Suppl. Fig. 1). In this group, COVID-19 severity increased with age (Suppl. figure. 2B) and anti-SARS-CoV-2 S1 IgG ratio increased in association with COVID-19 disease severity score (Suppl. figure 2C).

Among participants of the CoCo 1.0 cohort, however, not one of the positive baseline results of the anti-SARS-CoV-2 S1 IgG was confirmed independently and no sample scored subsequently positive in two assays (Figure 3). The cumulative incidence of cases with positive and borderline anti-SARS-CoV-2 S1 IgG results until week six of the study was 1·86% and 0·93%, respectively. To account for inter-assay variability among weekly ELISAs in the CoCo 1.0 cohort and to better identify ELISA results reflecting true seroconversion, all sera (from baseline to week six) of the eight HCP with positive or equivocal anti-SARS-CoV-2 S1 IgG/IgA ELISA results were reanalysed on single ELISA plate (Figure 4). Optical density ratios remained mostly stable, but some subjects showed declines in IgG ratios by more than 30% over 2-3 weeks (HCP 3 and 6) and others displayed oscillations in positive IgA ratios (HCP 2 and 5). Remarkably, only one subject (HCP1) clearly seroconverted for both anti-SARS-CoV-2 (S1) IgG and IgA during the study period with ELISA results turning from negative to increasingly positive. Samples of HCP1 obtained at week 1, 2, and 4 also turned positive in the anti-SARS-CoV-2 NCP IgG ELSIA and SARS-CoV-2 antibody rapid test (Figure 4). HCP1 had returned from Austria prior enrolment, but reported no signs and symptoms suggestive of SARS-CoV-2 infection during the entire study period.

**Figure 3:**
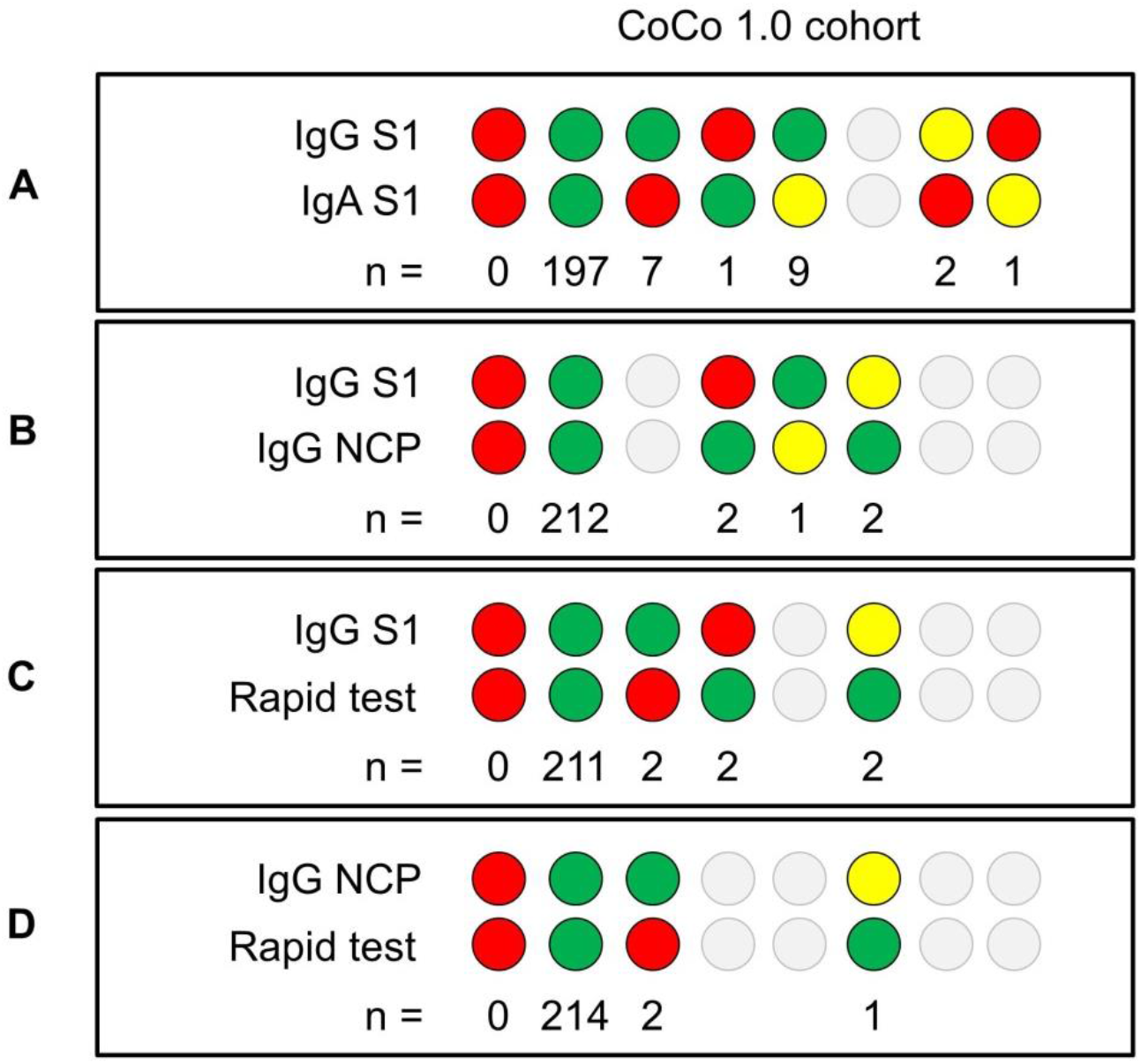
Consistency of seropositivity rates of the different serological testing systems applied in CoCo cohort 1.0. Results of the anti-SARS-CoV-2 S1 IgG versus IgA ELISA (**A**), anti-SARS-CoV-2 S1 IgG versus anti-SARS-CoV-2 NCP IgG ELISA (**B**), anti-SARS-CoV-2 S1 IgG ELISA versus the WANTAI anti-SARS-CoV-2 antibody rapid test (**C**), and anti-SARS-CoV-2 NCP IgG ELISA versus WANTAI anti-SARS-CoV-2 antibody rapid test (**D**). Red dots represent positive results (IgG ratio >1·1, positive band, respectively), yellow dots represent borderline positive results (IgG ratio 0·8-1·1), and green dots represent negative results (IgG ratio <0·8, no band, respectively).

**Figure 4:**
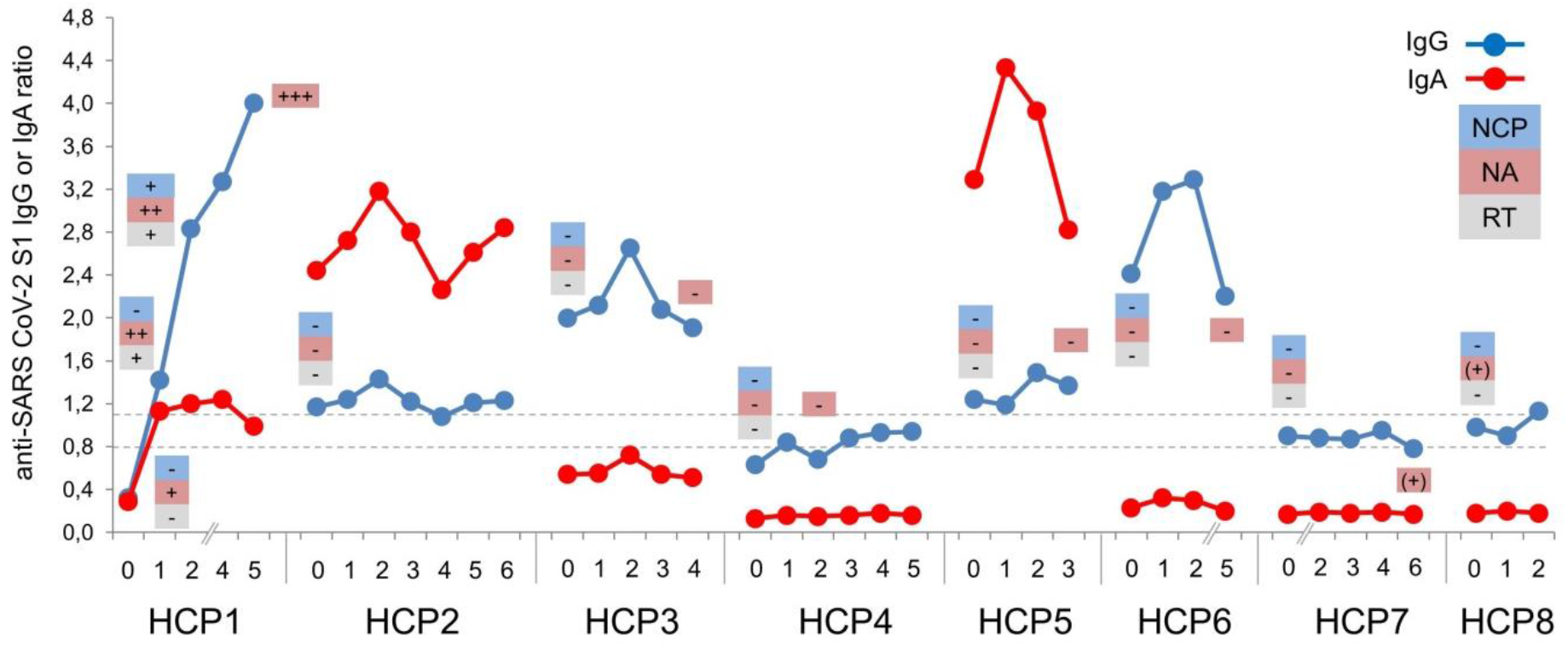
Serology results of eight HCP (1-8) in the CoCo cohort 1.0 with at least one positive or borderline positive anti-SARS-CoV-2 S1 IgG ELISA during the observation period. All samples from HCP with at least one positive or borderline result at any time point (HCP 1-8) were measured on one ELISA plate. Anti-SARS-CoV-2 S1 IgA is depicted in red, anti-SARS-CoV-2 S1 IgG depicted in blue. Results of anti-SARS-CoV-2 NCP (NCP), neutralisation assay (NA), or SARS-CoV-2 antibody rapid test (RT) from selected samples are indicated as positive or negative. The results of the neutralisation assay at IC50 are given as 1:2 (+), 1:8 +, 1:32 and 1:64 ++, 1:512 +++.

To assess whether the obtained positive anti-SARS-CoV-2 S1 IgG serology results represented true immune responses resulting in virus neutralisation activity, we performed plaque-assays using VeroE6 cells and *in vitro*-propagated SARS-CoV-2. Anti-SARS-CoV-2 S1 IgG results of COVID-19 patients correlated well with NT90 neutralisation (Suppl. 3A). Except for HCP1, where longitudinal analysis was strongly suggestive of seroconversion, all HCP with positive or borderline positive anti-SARS-CoV-2 S1 IgG lacked significant neutralisation activity against SARS-CoV-2 (Figure 4). HCP7 and HCP8 displayed very low neutralisation (1:2) in one of the two samples tested. In contrast, HCP1 developed strong SARS-CoV-2 neutralisation activity (NT50, 1:512) confirming SARS-CoV2 immunity. Interestingly, NT50 neutralisation was detectable (1:8) before anti-SARS-CoV-2 S1 IgG and IgA results turned positive.

Neutralisation assays performed using a SARS-CoV-2 variant are widely considered the gold standard, but require significant resources, safety lab requirements, and expertise. To employ a more readily applicable *in vitro* system, which would allow large-scale neutralisation assessments, we took advantage of a newly-established surrogate viral neutralisation test (sVNT), which assesses the degree to which serum antibodies can interfere with the binding of SARS-CoV-2-S-RBD to ACE2 *in vitro* ^17^. First, we demonstrated correlation of the sVNT data with results obtained from representative COVID-19 control samples in the plaque assay (r=0·833 p<0.001, Suppl. Figure 3B). In addition, we found excellent correlation of sVNT results at 1:180 (r=0·873, p<0.001) and 1:520 (r=0·945, p<0.001) dilutions with the ELISA anti-SARS-CoV-2 S1 IgG results (suppl. Figure 3C-D). The sVNT consistently identified inhibition in sera obtained from PCR-confirmed COVID-19 cases, for which strong neutralisation was detected by plaque assays (Figure 5A). Similarly, the serum of HCP1 of the CoCo cohort 1.0 showed increasing neutralisation capacity over time in the sVNT assay analogous to the plaque assay results (Figure 5B). However, except for HCP7, which at another time point showed borderline (NT50 1:20) neutralization signal (Figure 4), all HCP with positive or borderline anti-SARS-CoV-2 S1 IgG results had no evidence for inhibition in the sVNT (Figure 5C).

**Figure 5:**
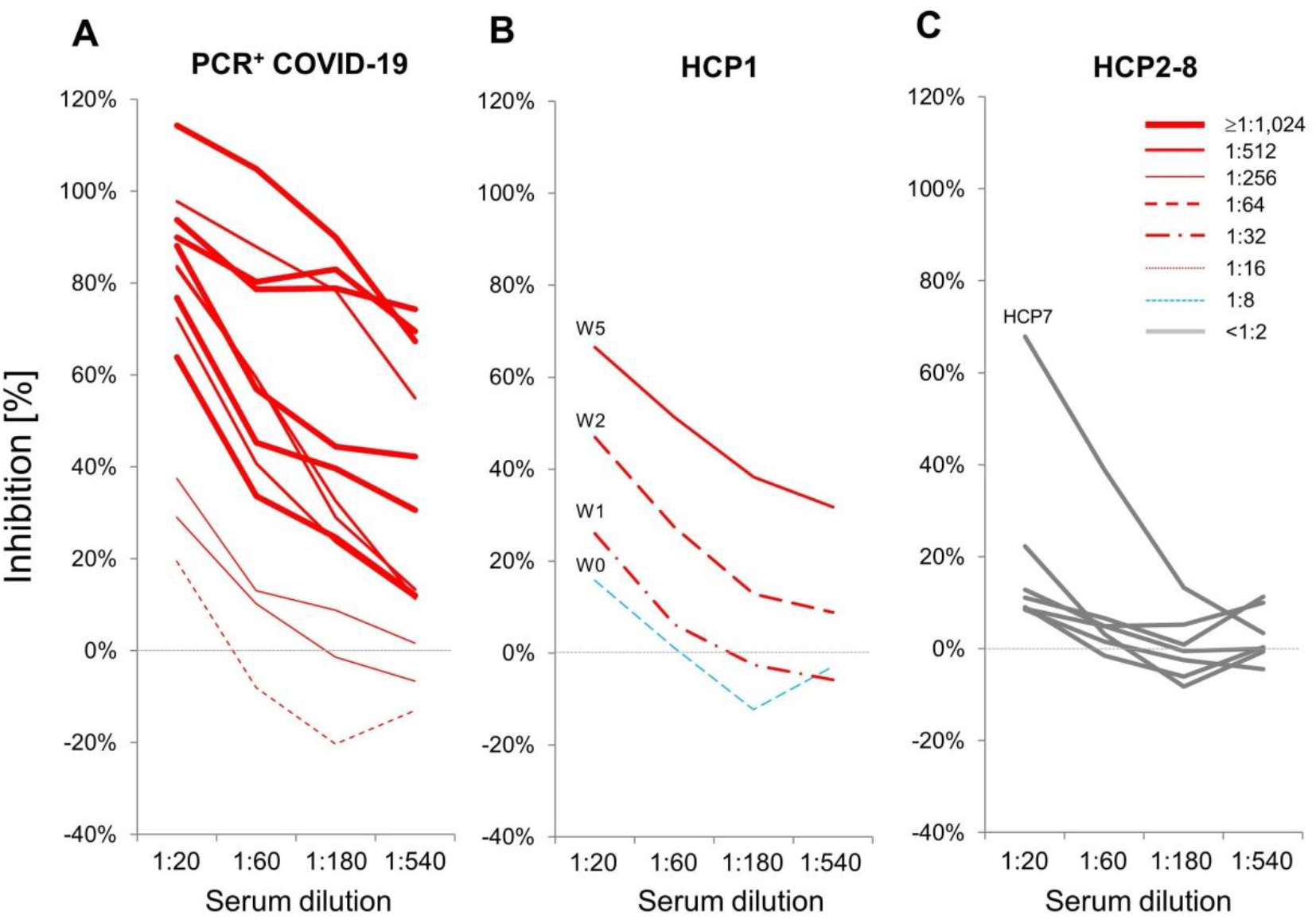
Inhibition in the sVNT compared to neutralisation activity in the plaque assay. (**A**) Sera of convalescent n=13 PCR^+^ COVID-19 patients with various neutralisation activity in the plaque assay (IC50, 1:16 to ≥ 1:1,024, red lines as indicated in the legend) are depicted in according to their percent inhibition activity in the sVNT at various dilutions as indicated. (**B**) Increase of inhibition in the sVNT during seroconversion [Week (W) 1 to 5] of HCP1 and rise in neutralisation activity in the plaques assay as depicted by the lines. (**C**) Inhibition results obtained in the sVNT with sera from HCP 2 to 8, which had least one positive or equivocal positive anti-SARS-CoV-2 S1 IgG ELISA result. None of these sera revealed significant neutralisation activity in the plaque assay (IC50 ≤1:2).

## Discussion

To the best of our knowledge, this is the first prospective longitudinal study on SARS-CoV-2 functional seroconversion and self-perceived infection risk in frontline HCP. We show that in a country with comparably low COVID-19 prevalence and an advanced, resource-rich health care system, the current rate of anti-SARS-CoV-2-Ig seroconversion in HCP after the peak of the pandemic is low, while the frequency of reported respiratory symptoms and the self-perceived risk for having contracted COVID-19 is considerable.

Since the emergence of the virus in late 2019 in China, the imminent threat to HCP of contracting COVID-19 in the workplace has been a pressing issue ^18^. High rates of asymptomatic infections, ranging from 18 to 88% ^6,19,20^, and transmissions before the onset of symptoms ^21^ have raised concerns about a potentially high rate of undiagnosed SARS-CoV-2 infections. In this context, serology studies are important to characterise transmission rates, and provide insight into humoral immunity to SARS-CoV-2.

Our study addresses several central questions about SARS-CoV-2 transmission and serological monitoring which are relevant to regions around the globe. In areas with resource-limited health care infrastructure and/or infection control, the pandemic has overwhelmed HCP and health care systems, recapitulating the initial wave of infections in Wuhan or the Lombardy region ^22^. However, other countries, such as Germany, were in the fortunate position of successfully flattening the exponential spread of the virus and were able to provide hospitals and care givers with appropriate PPE in a relatively timely fashion. In spite of this, a recent national survey collecting data during the time period covered by our study reported that over 60% of German HCP, particularly women, had concerns regarding their own health while working during the current pandemic ^23^. This observation is in line with our finding on self-perceived risks of having contracted SARS-CoV-2, particularly in female participants. Study participants were able to access their test results, which, as reported here, were overwhelmingly negative. This presumably contributed to the significant decline in risk perception over the study period. The high follow up rate in the CoCo 1.0 cohort, however, supports the idea that study participants were highly interested in their personal serological status.

The observed low cumulative incidence (0·46%) for functional anti-SARS-CoV-2 IgG is based on the combination of commercially available serology assays and in-house neutralisation tests and stands in contrast to reports from Spain, Italy, and UK, in which much higher seroconversion rates in HCP are reported ^2,10,24^. In addition to the low regional prevalence of COVID-19, sufficient access and rigorous use of PPE are likely to have contributed to this outcome. The only participant with confirmed seroconversion was most likely infected in a COVID-19 hotspot outside Germany. Interestingly, evidence for differences in the rate of hospital-acquired versus community-acquired SARS-CoV-2 infections is limited even in areas with higher COVID-19 disease burden ^2,8^.

Our data highlight the need for a cautious approach to serology screening and result interpretation in regions with low SARS-CoV-2 infection rates. Mass testing of both symptomatic and asymptomatic HCP has been proposed to reduce spread in mild or asymptomatic cases and to protect the health-care workforce ^25^. Nevertheless, given the differences in local spread dynamics, pre-testing probabilities and targeted screening approaches must be considered ^26^. The high rate and lack of discriminative value of respiratory symptoms observed in our cohort is consistent with findings from other groups ^27,28^, which show that non-respiratory symptoms in HCP (fever, anosmia/ageusia, muscle ache, ocular pain, general malaise and extreme tiredness) were associated with positive SARS-CoV-2 PCR results, while respiratory symptoms were not. Thus, non-respiratory symptoms are likely better measures of pre-test probability in symptomatic individuals.

Our results suggest that all positive results obtained by an ELISA tests from asymptomatic individuals or those with mild or unspecific symptoms should be confirmed or disproved by an alternative serology test. Whether this secondary, confirmatory testing should target different SARS-CoV-2 antigens or whether simply employing an alternate technique might suffice (e.g. chromatographic lateral flow rapid testing) will require further investigation. Alternatively, screening strategies may include parallel detection of immunoglobulins against human coronaviruses (HKU1, OC43, NL63, and 229E) with high potential for cross-reactivity.

Studies in patients with COVID-19, ranging from mildly symptomatic to critically ill, have consistently shown that almost all patients have detectable antibodies by day 28 ^25,29^. All our PCR+ COVID-19 study participants with negative results for anti-SARS-CoV-2 S1 IgG were tested at least 26 days after disease onset. Interestingly, all were females with mild disease. These factors have been suggested to be associated with weaker humoral anti-SARS-CoV-2 immunity. If illness severity correlates with anti-SARS-CoV-2 IgG responses and neutralisation potency ^14^, we hypothesise that asymptomatic SARS-CoV-2 infection could lead to a considerable number of transient “viral carriers” with undetectable systemic humoral immunity. These individuals would be missed by studies using serology screening only ^30,31^. The extent to which asymptomatic cases contribute to the current pandemic is currently unknown, and anti–SARS-CoV-2 responses in subclinical infections, as we demonstrate here, must be carefully characterised to better assess the rate of serological non-responders. The extent to which serological data can be employed to identify previously infected pauci- or asymptomatic persons remains unknown (5). Of note, cellular immunity against SARS-CoV-2 alone may confer protective immunity in the absence on antibody response ^32^.

A disadvantage of functional neutralisation assays is that they can only be performed by experienced staff in a biosafety level 3 laboratory due to the need to culture live virus. The surrogate neutralisation assay we use in this study has shown close correlation when compared to assays using live pseudotyped vesicular stomatitis virus (VSV) incorporating the S protein of SARS-CoV-2 ^17^ This assay consistently gave results analogous to the neutralisation assay with live SARS-CoV-2. This may become a useful tool for ascertaining robust systemic humoral immunity and assessing the kinetics of protective immunity.

Our study has several limitations. We did not perform molecular testing on respiratory specimens, which would provide information on viral carrier status in pauci- or asymptomatic HCP. We did not investigate localised immune responses, e.g. IgA in tears or mucosa fluids, or innate and cellular immune responses resulting from SARS-CoV-2 infection. Our questionnaire primarily focused on respiratory symptoms, which turned out to be of little discriminative value for identifying COVID-19. Our assessment of absolute self-perceived risk is probably a rough estimate, likely reflecting a composition of public and individual risk perception. Of note, this report represents an interim analysis, and the ongoing CoCo cohort 2.0 will likely provide more information on these topics.

In summary, our data show a low functional seroconversion rate in HCP, contrasting with a considerable self-perceived infection probability. Self-reported respiratory symptoms appear to be too unspecific to inform pre-test probability and serology test result interpretation. Our data highlight the need for a cautious approach to serology screening and result interpretation in regions with low SARS-CoV-2 infection rates. For analyses of humoral SARS-CoV-2 specific immune response in a low pre-test probability setting, positive results from single measurements should be confirmed by alternative serology tests or functional assays.

## Data Availability

The datasets used and/or analysed during the current study are available from the corresponding author on reasonable request.

## Ethics approval and consent to participate

The here presented analyses were approved by local authorities (Data Security Management and Institutional Review Board of Hannover Medical School), approval number 8973_BO_K_2020). Informed consent was obtained from all participants.

## Consent for publication

Not applicable

## Acknowledgement

We thank Marion Hitzigrath, Melanie Ignacio, Annkathrin Anton, Till Redeker, Madeleine Rommel, Luis Manthey, and Nele Stein for technical and logistical support. We thank the CoCo Study team for help with enrolment and blood collection for the Coco 1.0 Cohort(Martina Toussaint, Marcel Winkelmann, Mark Greer, Marcus Bachmann, and Birgit Heinisch) and all HCP of the CoCo Study for their participation. This study is supported by an unrestricted grant from Novartis to GMNB, AJ, DE, CH, from PARI to CH, AJ, CD, MW, and Verein Kinderherz Hannover. GMNB, MW and AJ are supported by the German Centre for Infection Research (DZIF). RF is supported by Deutsche Forschungsgemeinschaft, DFG Excellence Strategy - EXC 2155 “RESIST”- project number 39087428, Sonderforschungsbereich SFB900 project number 158989968 - SFB 900 and by a fund of the state of Lower Saxony (14 - 76103-184 CORONA-11/20). The funders had no role in study design, data collection and analysis, decision to publish, or preparation of the manuscript.

## Author’s contribution

GMNB, CH, AJ designed and managed the project, analysed the data and drafted the manuscript. AC managed study enrolment, designed the workflow and questionnaires, organised data collection, and biobanking and analysed the data. MS performed the ELISA and rapid test experiments and designed the laboratory workflow. RF and BB developed the sVNT and BB and SW performed the inhibition experiments; HS and BS performed and analysed the neutralisation assays. TT, TG, MZK, A-LB, ATT, AZS, AH, CD, MW, IP, TW, and DE supported recruitment, blood sampling and follow up visits. All authors discussed the results and approved the final manuscript.

## Supplementary information

(Cossmann A, et al. CoCo Study)

### Methods

**Study design, enrolment, and follow up:** Study participants of the CoCo cohort 1.0 were enrolled between March 23rd and April 17^th^ 2020, at the peak of the SARS-CoV-2 pandemic in Germany. Week 0 of the study was March 23rd - March 29th. n=217 HCP from clinical units involved in COVID-19 patient care were eligible (emergency department: n=58 HCP, infectious and pulmonary disease wards: n=28 HCP, pediatric departments: n=44 HCP, neurology department: n=58 HCP, rheumatology and immunology department: n=19 HCP, trauma surgery: n=10 HCP). No incentives were offered upon enrollment, but participants had access to their ELISA results via web-based personalised access codes. The questionnaire assessed typical respiratory symptoms also reported by COVID-19 patient as follows: cough, fever, dyspnea, rhinitis, sore throat, and loss of sense of taste and smell. Blood samples were either drawn by study participants themselves or by members of the research team, and serum was prepared and stored for analysis at −80°C. Anti-SARS-CoV-2 S1 IgG ELISA testing was performed in weekly batches. Enrollment for CoCo 2.0 cohort began in May 2020 with the goal of recruiting a significantly larger sample of HCP from a wide variety of departments in out hospital for serological testing every six months.

To further analyse serological responses in a variety of COVID-19 manifestations and severities, blood samples were obtained from n=41 polymerase chain reaction (PCR) -confirmed convalescent COVID-19 patients treated either at Hanover Medical School or by general practitioners in the region of Hannover as positive controls (n=21 male, n=20 female, at least 10 days post recovery, mean time since start of symptoms 35·3 days, range 11-70 days, mild disease: n=29, moderate disease: n=6, severe disease: n=9). Blood from these patients was drawn ≥10 days after symptom onset and all patients gave written informed consent. Eleven samples and associated data were provided by the Hanover Unified Biobank (HUB), the central biobank of Hanover Medical School (MHH) in accordance with the regulations of HUB and the approval of the ethics committee. Clinical data (e.g. disease score: “mild” = out-patient, “moderate” = regular in-patient unit or infectious disease unit, “severe” = ICU and mechanic ventilation, symptoms/duration of symptoms) were derived from hospital data files or questionnaire-based self-reporting.

**Laboratory testing:** Baseline antibody testing results for anti-SARS-CoV-2 S1 IgG and IgA of the CoCo cohort 1.0 were previously reported (16). As a primary testing system, a semiquantitative ELISA based on the SARS-COV-2 S1 spike protein domain/receptor binding domain (Euroimmun, Lübeck, Germany - CE certified version: specificity 99.0 %, sensitivity 93·8 % after day 20 according to the manufacturer) was used for weekly measurements. We previously confirmed specificity for SARS-CoV-2 IgG in this assay with 99·3% (16) and used the same historic controls (n=156 sera from non-European refugees and migrants from 2015) to calculate a specificity of 97·5% for SARS-CoV-2 IgA. For additional retrospective analyses in all baseline samples, positive controls, and positive or equivocal positive sera from the above mentioned anti-SARS-CoV-2 S1 IgG screening, two additional tests were performed: First, a recently available anti-SARS-CoV-2-nucleocapsid protein (NCP) IgG ELISA based on a modified NCP (Euroimmun, Lübeck, Germany - CE certified version: specificity 99·8 %, sensitivity 94·6 % after day 10 according to manufacturer) and second, we also tested the WANTAI SARS-CoV-2 antibody rapid test (SZABO SCANDIC, Vienna, Austria - CE certified version), which employs a chromatographic lateral flow device in a cassette format. Colloidal gold conjugated recombinant antigens corresponding to SARS-CoV-2 are dry-immobilised at the end of a nitrocellulose membrane strip. To test our study subjects, 10μl serum was placed in the specimen window and procedures were performed according to the manufacturer’s instructions. If present in the specimen, SARS-CoV-2 antibodies bind to the gold conjugated antigens and form particles, and are captured by the SARS-CoV-2 antibody generating a visible red line. Results were read by two independent investigators and documented by photography. The data sheet (Feb 26^th^, 2020) reports a sensitivity of 95·6% (131/137) and a specificity of 95·2% (199/209).

**Neutralization assay:** Neutralization assays were performed using an in vitro-propagated SARS-CoV-2 strain isolated in Bonn/Germany via nasopharyngeal swabbing of a patient from Heinsberg, Germany. To test SARS-CoV-2 neutralization capacities, serum samples from study participants were inactivated at 56°C for 30 min and insoluble matter was pelleted by centrifugation at 3,000 rpm for 5 min. The supernatant was transferred to a new tube, mixed and used in a plaque reduction neutralization assay. To this end, heat inactivated plasma samples were serially prediluted in OptiPROTMSFM (Gibco) starting with 1:2 up to 1:1,024. 120 μl of each plasma dilution was mixed with 80 plaque forming units (PFU) of SARS-CoV-2 in 120 μl OptiPROTMSFM. After 1 h incubation at 37°C, 200 μl of each mixture was added to wells of a 24-well plate seeded the day before at 1.25×10^5^ Vero E6 cells/well. After incubation for 1 h at 37°C, the inoculum was removed and cells were overlayed with a 1:1 mixture of 1.5% (w/v) carboxymethylcellulose (Sigma) and 2xMEM (Biochrom) with 4% (v/v) FBS (Gibco), 4.4 g/L NaHCO_3_ and 200 U/ml Penicillin/Streptomycin. After incubation at 37°C and 5% CO2 for three days, the overlay was removed and 24-well plates were fixed using a 6% (v/v) formaldehyde solution, rinsed with H_2_O and stained with 1% (w/v) crystal violet (Merck) in 50% (v/v) ethanol. Neutralizing titers were calculated as the reciprocal of serum dilutions resulting in neutralization of 50% or 90% input virus (NT50/NT90, respectively), read out as reduction in the number of plaques.

**Supplementary figure 1:**
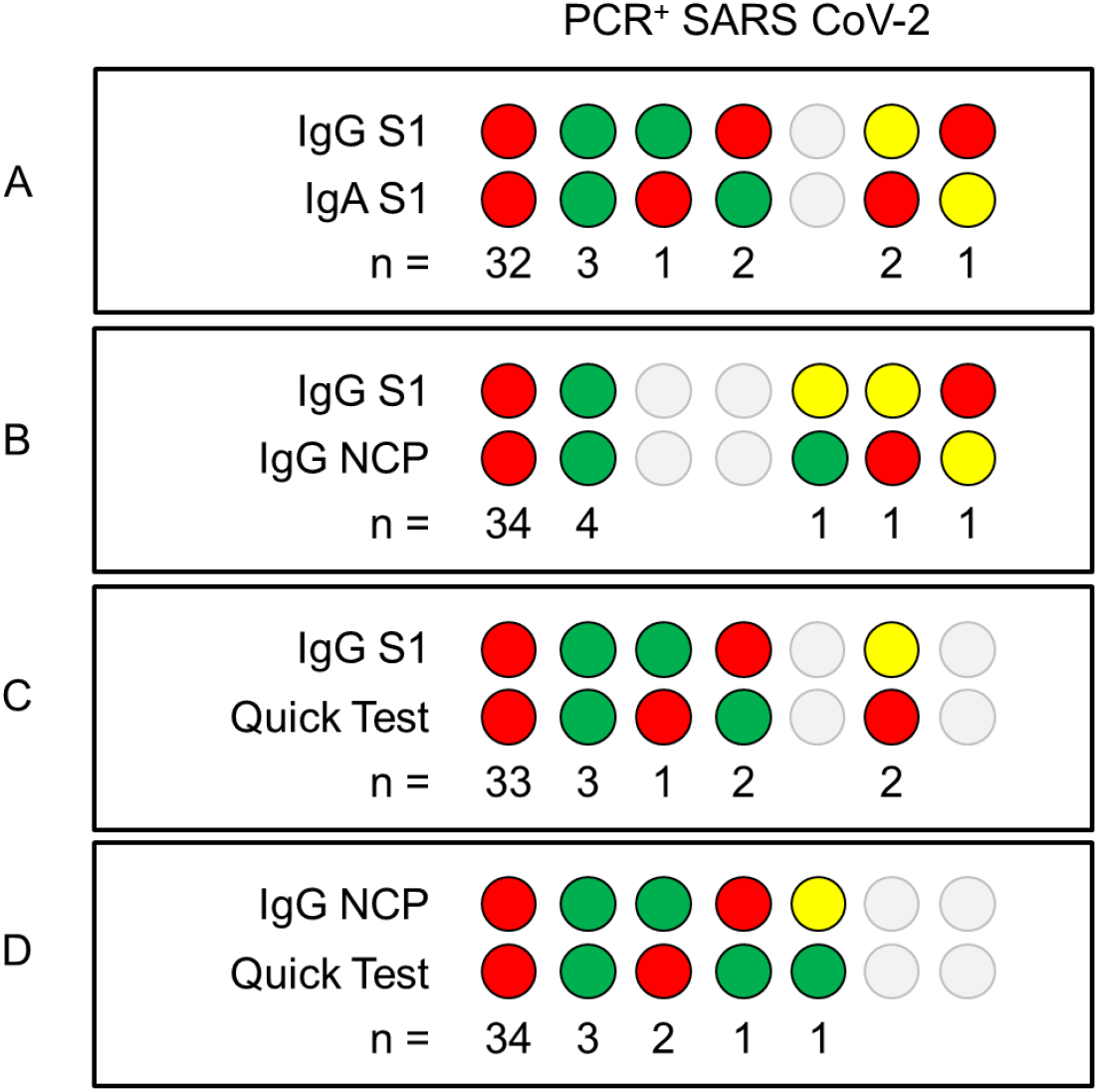
Serology results of the PCR+ COVID-19 patients. To assess the sensitivity, specificity, and concordance of the different serological testing systems, we compared the performance of the different assays in a set of positive controls (n=41 adult patients collected after recovery of PCR-confirmed COVID-19, 51% male, mean age 45·3+2·5 years; mean duration of symptoms 18·3+2·2 days, range 3-50 days; mean time since start of symptoms 35·3 days, range 11-70 days). (**A-D**) depict the individual results of the serological tests as indicated. Red dots represent positive results (IgG ratio >1.1, positive band, respectively), yellow dots represent equivocal positive results (IgG ratio 0.8-1.1), and green dots represent negative results (IgG ratio <0.8, no band, respectively). (**A**) N=35/41 tested positive (plus n=2 equivocal positive) for anti-SARS-CoV-2 S1 IgG (sensitivity 87.2%) and n=34/41 were positive for anti-SARS-CoV-2 S1 IgA (sensitivity 85 4%). Consistent results for anti-SARS-CoV-2 IgG and IgA were observed in n=35 out of 41 COVID-19 patients (77.7%). (**B**) Overall, the anti-SARS-CoV-2 S1 IgG and anti-SARS-CoV-2 NCP IgG ELISAs gave most consistent results (38/41 with similar results). However, both S1 and NCP tests combined identified no more than n=35/41samples as clearly positive (sensitivity 87·5%). (**C**) Best sensitivity (85·7%) was demonstrated by the SARS-CoV-2 antibody rapid test, which detected 36/41 as positive.

**Supplementary figure 2:**
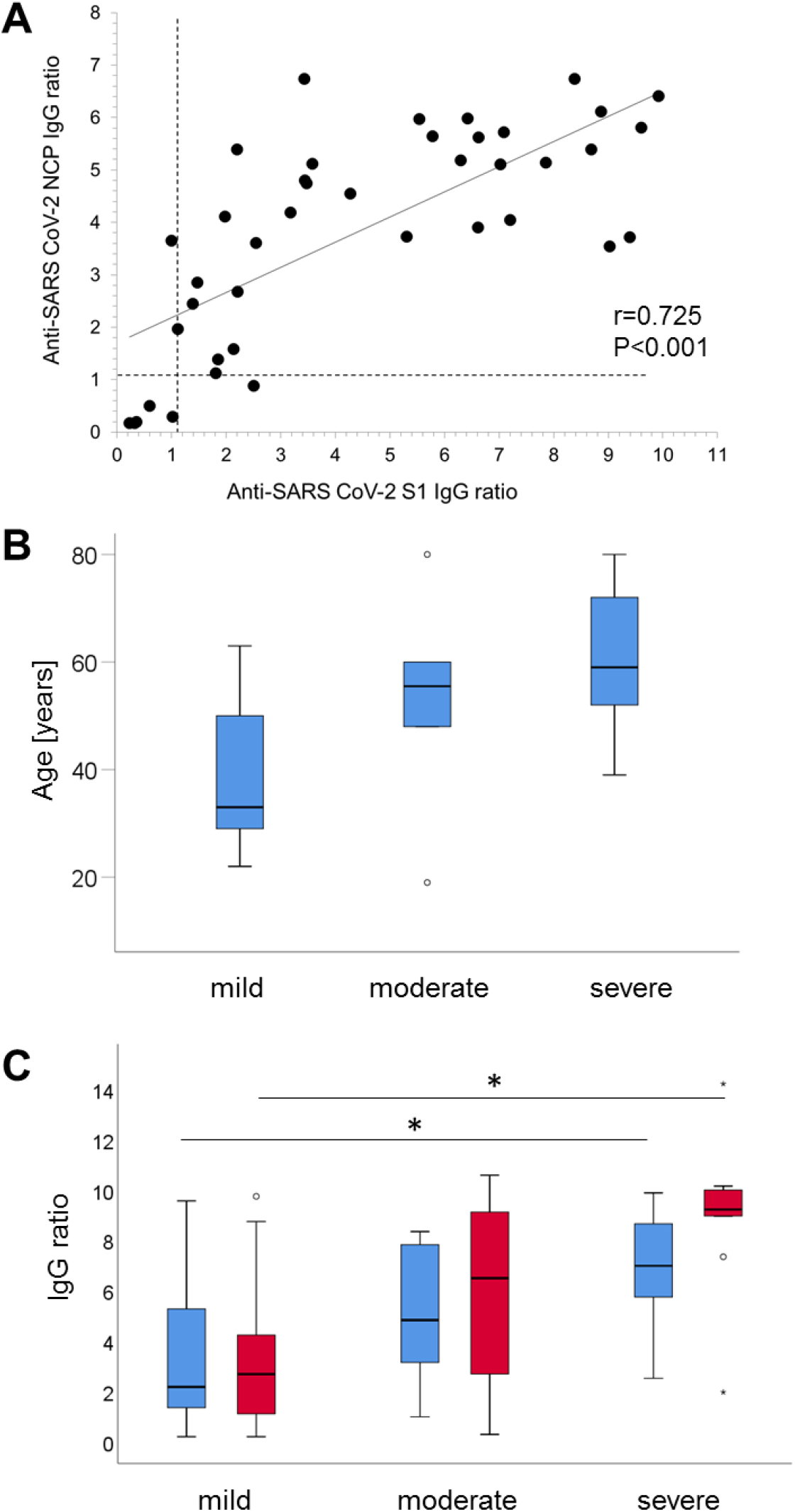
Association of disease activity scores to ELISA results and age in PCR+ COVID-19 patients. (**A**) Correlation of anti-SARS-CoV-2 S1 IgG and anti-SARS-CoV-2 NCP IgG results from n=41 PCR-confirmed COVID-19 patients. The dotted line indicates the cut off for positivity according to the manufacture’s recommendation (IgG ratio >11). (**B**) The level of anti-SARS-CoV-2 S1 IgG ELISA results (IgG ratio in blue and IgA ratio in red) is depicted in relation to the age of COVID-19 patients and (**C**) to a three grade COVID-19 severity score (* p<0 05).

**Supplementary figure 3:**
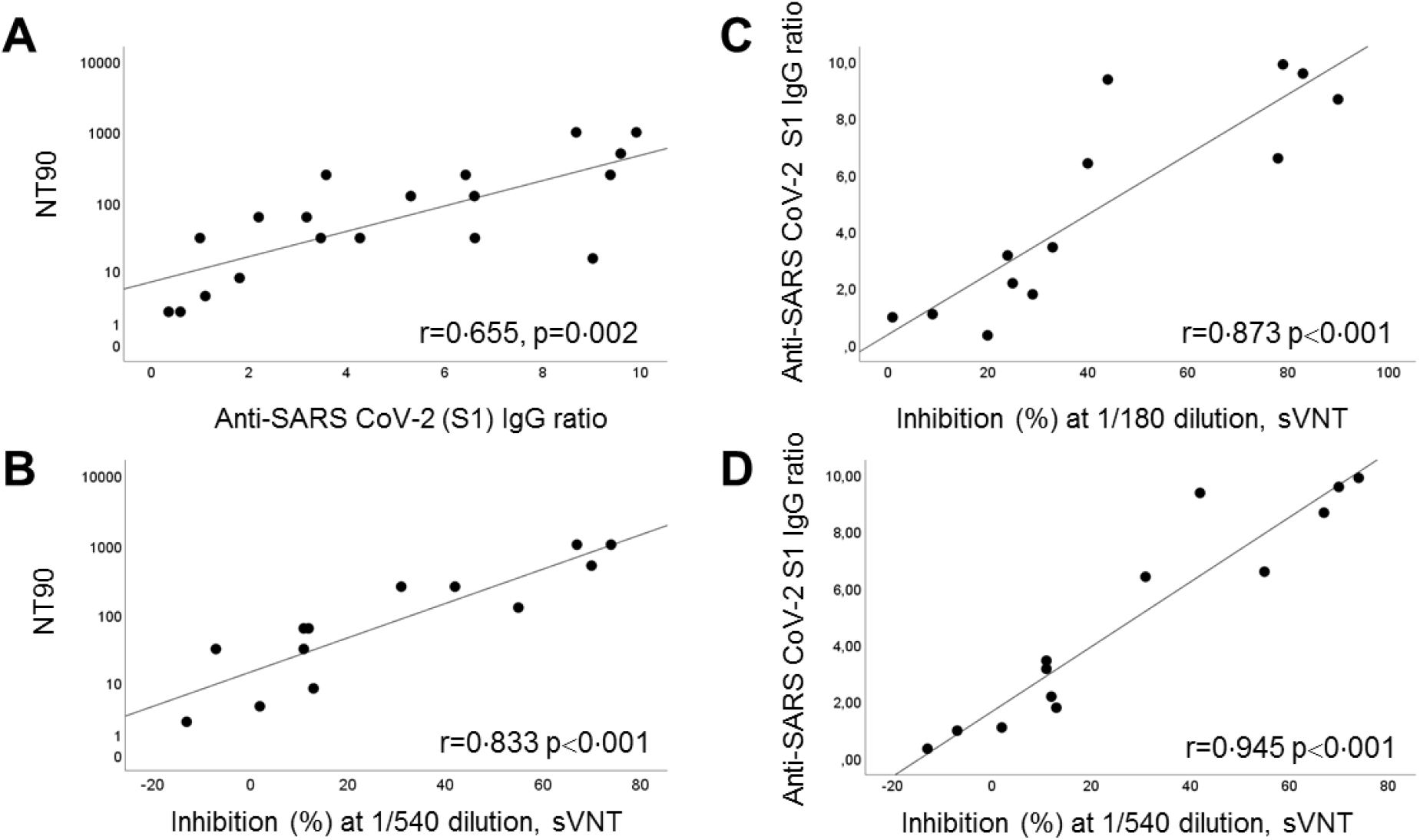
Correlation of serology, inhibition in the sVNT, and neutralisation activity in sera from PCR+ COVID-19 patients. In (**A**) the neutralisation titer (NT90) in relation to the anti-SARS-CoV-2 S1 IgG ratio is depicted (n=19). (**B**) shows the correlation of the neutralisation (NT90) and the percent inhibition in the sVNT at 1:540 dilution (n=13). Panel **C** and **D** represent the correlation of the anti-SARS-CoV-2 S1 IgG ratio results to the inhibition in the sVNT at dilutions of 1:180 and 1:540 (n=13).

